# SARS-CoV-2 serological analysis of COVID-19 hospitalized patients, pauci-symptomatic individuals and blood donors

**DOI:** 10.1101/2020.04.21.20068858

**Authors:** Ludivine Grzelak, Sarah Temmam, Cyril Planchais, Caroline Demeret, Christèle Huon, Florence Guivel-Benhassine, Isabelle Staropoli, Maxime Chazal, Jeremy Dufloo, Delphine Planas, Julian Buchrieser, Maaran Michael Rajah, Remy Robinot, Françoise Porrot, Mélanie Albert, Kuang-Yu Chen, Bernadette Crescenzo, Flora Donati, François Anna, Philippe Souque, Marion Gransagne, Jacques Bellalou, Mireille Nowakowski, Marija Backovic, Lila Bouadma, Lucie Le Fevre, Quentin Le Hingrat, Diane Descamps, Annabelle Pourbaix, Yazdan Yazdanpanah, Laura Tondeur, Camille Besombes, Marie-Noëlle Ungeheuer, Guillaume Mellon, Pascal Morel, Simon Rolland, Felix Rey, Sylvie Behillil, Vincent Enouf, Audrey Lemaitre, Marie-Aude Créach, Stephane Petres, Nicolas Escriou, Pierre Charneau, Arnaud Fontanet, Bruno Hoen, Timothée Bruel, Marc Eloit, Hugo Mouquet, Olivier Schwartz, Sylvie van der Werf

## Abstract

It is of paramount importance to evaluate the prevalence of both asymptomatic and symptomatic cases of SARS-CoV-2 infection and their antibody response profile. Here, we performed a pilot study to assess the levels of anti-SARS-CoV-2 antibodies in samples taken from 491 pre-epidemic individuals, 51 patients from Hôpital Bichat (Paris), 209 pauci-symptomatic individuals in the French Oise region and 200 contemporary Oise blood donors. Two in-house ELISA assays, that recognize the full-length nucleoprotein (N) or trimeric Spike (S) ectodomain were implemented. We also developed two novel assays: the S-Flow assay, which is based on the recognition of S at the cell surface by flow-cytometry, and the LIPS assay that recognizes diverse antigens (including S1 or N C-terminal domain) by immunoprecipitation. Overall, the results obtained with the four assays were similar, with differences in sensitivity that can be attributed to the technique and the antigen in use. High antibody titers were associated with neutralisation activity, assessed using infectious SARS-CoV-2 or lentiviral-S pseudotypes. In hospitalized patients, seroconversion and neutralisation occurred on 5-14 days post symptom onset, confirming previous studies. Seropositivity was detected in 29% of pauci-symptomatic individuals within 15 days post-symptoms and 3 % of blood of healthy donors collected in the area of a cluster of COVID cases. Altogether, our assays allow for a broad evaluation of SARS-CoV2 seroprevalence and antibody profiling in different population subsets.

## Introduction

About four months after the initial description of atypical pneumonia cases in Wuhan in December 2019, COVID-19 has become a major pandemic threat. As of April 14, 2020, about half of the human population is under confinement, 2 million infections have been officially diagnosed, with 121,000 fatalities and 0.5 million recovered cases. COVID-19 is caused by SARS-CoV-2 ^1 2^, a betacoronavirus displaying 80% nucleotide homology with Severe Acute Respiratory Syndrome virus (now termed SARS-CoV-1), that was responsible for an outbreak of 8,000 estimated cases in 2003.

PCR-based tests are widely used for COVID-19 diagnosis and for detection and quantification of SARS-CoV2 RNA ^3 4 5^. These virological assays are instrumental to monitor individuals with active infections. The average virus RNA load is 10^5^ copies per nasal or oropharyngeal swab at day 5 post symptom onsets and may reach 10^8^ copies ^6^. A decline occurs after days 10-11, but viral RNA can be detected up to day 28 post-onset in recovered patients at a time when antibodies (Abs) are most often readily detectable ^6 7^. Disease severity correlates with viral loads, and elderly patients, who are particularly sensitive to infection, display higher viral loads ^6 7^.

Serological assays are also being implemented. Anti-Spike (S) and Nucleoprotein (N) humoral responses in COVID-19 patients are assessed, because the two proteins are highly immunogenic. The viral spike (S) protein allows viral binding and entry into target cells. S binding to a cellular receptor, angiotensin-converting enzyme 2 (ACE2) for SARS-CoV-1 and -CoV2, is followed by S cleavage and priming by the cellular protease TMPRSS2 or other endosomal proteases ^8^. S genes from SARS-CoV and -CoV2 share 76% amino-acid similarity ^2^. One noticeable difference between the two viruses is the presence of a furin cleavage site in SARS-CoV2, which is suspected to enhance viral infectivity ^2^. The structures of S from SARS-CoV-1 and Co-V-2 in complex with ACE2 have been elucidated ^9-11^. S consists of three S1-S2 dimers, displaying different conformational changes upon virus entry leading to fusion ^9,10,12^. Some anti-S antibodies, including those targeting the receptor binding domain (RBD), display a neutralizing activity, but their relative frequency among the generated anti-SARS-CoV-2 antibodies during infection remains poorly characterized. The nucleoprotein N is highly conserved between SARS-CoV1 and -CoV2 (96% amino-acid homology). N plays a crucial role in subgenomic viral RNA transcription and viral replication and assembly.

Serological assays are currently being performed using in-house, pre-commercial versions or commercially available ELISA-based diagnostics tests ^6,7,13-15^. Other techniques, including point-of-care and auto-tests are also becoming available. In hospitalized patients, seroconversion is typically detected between 5-14 days post symptom onset, with a median time of 5-12 days for anti-S IgM and 14 days for IgG and IgA ^6,7,13-16^. The kinetics of anti-N response was described to be similar to that of anti-S, although N responses might appear earlier ^15-17^. Anti-SARS-CoV-2 antibody titers correlate with disease severity, likely reflecting higher viral replication rates and/or immune activation in patients with severe outcome. Besides N and S, antibody responses to other viral proteins (mainly ORF9b and NSP5) were also identified by antibody microarray ^17^.

Neutralisation titers observed in individuals infected with other coronaviruses, such as MERS-CoV, are considered to be relatively low ^6,18^. With SARS-CoV-2, neutralizing antibodies (Nabs) have been detected in symptomatic individuals ^6,8,19,20^, and their potency seems to be associated with high levels of antibodies. Neutralisation is assessed using plaque neutralisation assays, microneutralisation assays, or inhibition of infection with viral pseudotypes carrying the S protein ^6,8,19-21^. Of note, potent monoclonal NAbs that target RBD have been cloned from infected individuals ^22^. Whether asymptomatic infections, which are currently often undocumented ^23^, and most likely represent the majority of SARS-CoV-2 cases lead to protective immunity, and whether this immunity is mediated by NAbs, remain outstanding questions.

Commercial tests are not yet widely distributed. Thus, we have designed anti-N and anti-S ELISA, as well as novel assays for anti-SARS-CoV-2 antibody detection and neutralisation. We compared their performance and carried out anti-SARS-CoV-2 antibody profiling in different population subsets.

## Results

### Description of the serological tests

We first designed four tests to assess the levels of anti-SARS-CoV-2 antibodies in human sera. Their characteristics are summarized table 1.

**Table 1.** Serological assays used in the study

| Assay | Antigen | Serum dilution | Read-out |
| --- | --- | --- | --- |
| ELISA N | N | 1:200 | Optical Density |
| ELISA tri-S | Trimeric S | 1:400 | Optical Density |
| S-Flow | S at the cell surface | 1:300 | Flow cytometry |
| LIPS | S1 and N | 1:10 | Bioluminescence<br>(luciferase) |

### ELISAs

The two in-house ELISAs are classical tests, using as target antigens the full-length N protein (ELISA N) or the extracellular domain of S in a trimerized form (ELISA tri-S). The two recombinant antigens were produced in E. Coli (N) or in human cells (S).

The ELISA N assay is a classical indirect test for the detection of total immunoglobulins, using plates coated with a purified His-tagged SARS-CoV 2 N protein. Titration curves of sera from 22 COVID-19 patients and 4 pre-pandemic sera initially led to the determination that a dilution of 1:200 was of optimal sensitivity and specificity, and was later used for testing of large cohorts.

The ELISA tri-S, for trimeric S, allows for the detection of IgG antibodies directed against the SARS-CoV-2 Spike. We developed an ELISA using as antigen a purified, recombinant and tagged form of the S glycoprotein ectodomain, which was stabilized and trimerized using a foldon motif. Serum IgG from pre-epidemic (n=100), pauci-symptomatic (n=209), and hospitalized individuals (n=159) were titrated using serum dilutions ranging from 1:100 to 1:1,638,400 (Fig. S1). Receiving-operating characteristic analyses using either total area under the curve or single optical density measurements indicated that the 1:400 dilution provides the best sensitivity and specificity values and was therefore used in subsequent analyses (Fig. S1). Of note, the tri-S ELISA also permitted the titration of anti-S IgM and IgA antibodies in human sera (Fig. S1).

### S-Flow

The third assay, termed S-Flow, is based on the recognition of the S protein expressed at the surface of 293T cells (293T-S cells). We reasoned that in-situ expression of S will allow detection of antibodies binding to various conformations and domains of the viral glycoprotein. We verified that S was functionally active, by mixing 293T-S cells with target cells expressing ACE2. Large and numerous syncytia were detected, indicating that S binds to its receptor and performs fusion (not shown). The principle of the S-Flow assay is depicted Fig. S2A. 293T-S cells are incubated with dilutions of sera to be tested. Antibody binding is detected by adding a fluorescent secondary antibody (anti-IgG or anti-IgM). The signal is measured by flow-cytometry using an automated 96-well plate holder. The background signal is measured in 293T cells lacking S and subtracted in order to define a specific signal and a cut-off for positivity.

To establish the specificity of the assay, we first analyzed a series of 40 sera collected before 2019, from the Institut Pasteur biobank (ICAReB). All sera were negative (Fig. S2), strongly suggesting that antibodies against other coronaviruses circulating in France were not detected. We then measured the sensitivity of the assay, by assessing the reactivity of sera from Covid-19 patients hospitalized at Hôpital Bichat (Table S1). An example of binding with two patients’ sera (B1 and B2) is depicted Fig. S2B. Serial dilutions allowed for the determination of a titer, which reached a value of 24,600 and 2,700 for B1 and B2, respectively (Fig. S2B). Of note, the median fluorescence intensity (MFI) of the signal decreased with the dilution, indicating that MFI, in addition to the % of positive cells, provides a quantitative measurement of the levels of specific antibodies. We thus selected a single dilution (1:300) to analyze large numbers of samples. We then analyzed samples from 9 patients (B1-B9) (Fig. S2C and Table S1). We observed an increase of the IgG response over time, with positivity appearing 6 days after symptoms onset. Serial dilutions indicated that antibody titers raised over time (not shown). We observed similar patterns with the IgM and IgG responses (Fig. S2D). The absence of an earlier IgM response may be due to the lower sensitivity of the secondary anti-IgM antibodies tested or because of a short delay between the two responses, which has been already observed in COVID-19 patients. Addressing this question will require the analysis of a higher number of individuals. We also tested a secondary anti-whole Ig antibody, but it did not prove more sensitive than the anti-IgG. We thus tested the different cohorts with the secondary anti-IgG.

### LIPS

The fourth assay, termed LIPS (Luciferase Immunoprecipitation Assay) is based on the use of antigens made of viral proteins (or domains) fused to nanoluciferase (nanoluc) (Fig. S3). The objective was to develop an assay that is able to test large diverse cohorts and evaluate the range of antibody responses against a set of viral proteins or domains. This opens the possibility to select the best antigens for high throughput binding assays. Each antigen is used at the same molar concentration, based on a standardisation by luciferase activity of the amount of Ag engaged in each reaction. This allows for easy direct comparison of the Ab responses (amplitude and kinetic) against each antigen. A panel of 10 different S and N-derived antigens were first evaluated with a set of 34 pre-epidemic human sera were along with those of with 6 COVID hospitalized patients (Fig. S3). Two patients were sampled at 3 different time points. The strongest signals in COVID patients’ sera compared to background of pre-epidemic sera were identified with S1, S2 and N (C-term part) antigens (Fig. S3). Additional investigations on a limited panel of sera sampled in pauci-symptomatic patients showed that S2 responses were, regarding the diagnostic sensitivity and quantitative responses, similar to full S responses evaluated by S-Flow (Fig. S3). To avoid redundancy, we focused LIPS analysis to N, selecting it for its sensitivity regarding an intracellular viral protein not targeted by NAbs and S1 as it is described as a target of most NAbs. To establish the specificity of the assay, we first analyzed the same series of 40 sera we used for S-Flow and found all of the sera to be negative (Fig. S3). We also measured the kinetic of apparition of antibodies in the same longitudinal samples from 5 patients (Fig. 2 and Table 2). We observed an increase of response overtime, with positivity appearing 7-10 days after symptoms onset. Of note, the protein A/G beads used for precipitation of the immune complexes do not bind efficiently to IgM or IgA. Protein L, which has a higher affinity binding to IgA, has not yet been tested.

**Table 2.** Characteristics of the cohorts

| Cohorts | n | Samples | Date | Area | COVID-19 |
| --- | --- | --- | --- | --- | --- |
| pre-epidemic individuals | 491 | 491 | 2017-2019 | France | Naïve |
| hospitalized patients | 51 | 161 | Jan-March, 2020 | Paris<br>France | Confirmed |
| pauci-symptomatic individuals | 209 | 209 | March 3-4, 2020 | Crépy-en-Vallois,<br>France | Suspected |
| blood donors | 200 | 200 | March 20-24, 2020 | Lille, France | Unknown |

**Fig. 1:**
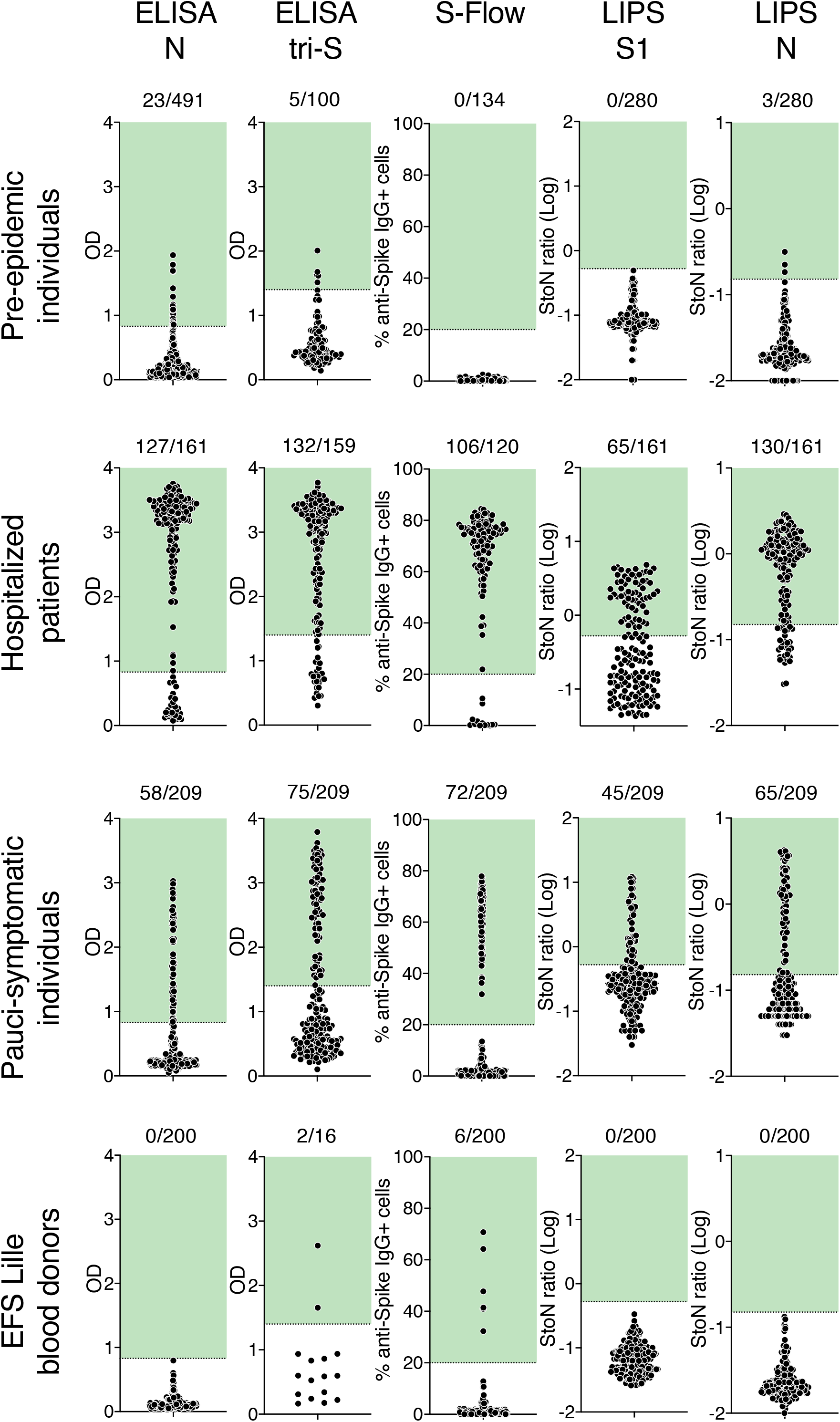
Serological survey of SARS-Cov-2 antibodies. Sera from pre-epidemic individuals sampled between 2017 and 2019 (first row), hospitalized cases with confirmed COVID-19 (second row), pauci-symptomatics individual from the Crépy-en-Vallois epidemic cluster (third row) and healthy blood donors (last row) were surveyed for anti-SARS-Cov-2 antibodies using four serological assays. ELISA N and ELISA tri-S are conventional ELISA, using either N or the trimeric ectodomain of S protein as antigens. S-Flow is an assay detecting antibodies bound to cells expressing S by flow cytometry. LIPS S1 and N detect either S1 or N fused to luciferase by immunoprecipitation. Pre-epidemic samples were used to determine the cut-off of each assay, which is indicated by a dashed line and a green area. ELISAs were set to 95% specificity. The number of positive samples is indicated. Each dot represents a sample.

**Fig. 2.**
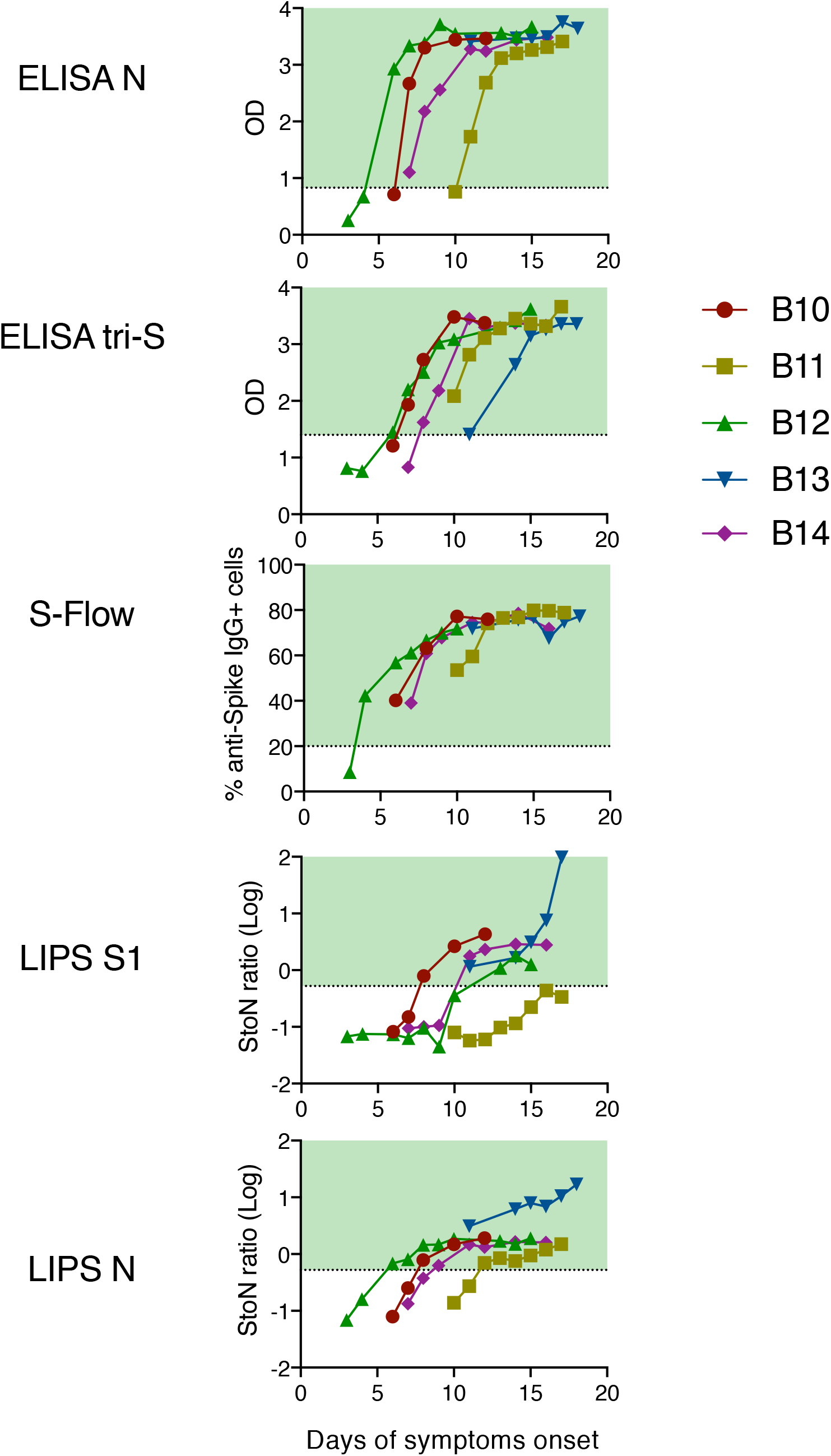
Antibody detection in 5 hospitalized patients. Kinetics of seroconversion in 5 hospitalized patients with at least 5 longitudinal samples. All patients were admitted in intensive care unit. Each line represents a participant. Dashed lines and green areas indicate assays cut-off of positivity.

### Description of the groups

We screened different cohorts to evaluate the performance of the four assays and corresponding antigens (Table 2). We first used sera from up to 491 pre-epidemic individuals, collected before 2019, to assess the specificity of the tests. We then measured antibody levels in 51 hospitalized COVID-19 patients from Hôpital Bichat (Paris), to determine the sensitivity of the tests and analyze the kinetics of seroconversion. The clinical and virological characteristics of four of these patients have been recently described ^24^. We next studied the prevalence of SARS-COV-2 positive individuals in a cohort of pauci-symptomatic individuals in Crepy-en-Valois, a city of 15,000 inhabitants in Oise. On 24 February 2020, a staff member from a high school in Crepy-en-Valois was admitted to an hospital in Paris with confirmed SARS-CoV-2 infection. On March 3-4, students from the high school, parents of the students, teachers and staff were invited to participate to an epidemiological investigation around this case. 209 blood samples were collected from individuals reporting mild signs compatible with COVID-19 (fever, cough or dyspnea). Finally, we tested 200 sera from blood donors from the Etablissement Français du Sang (EFS) in Lille (France). The blood samples were donated in two cities, Clermont (10,000 inhabitants) on March 20 and Noyon (13,000 inhabitants) on March 24, each located at about 60 kilometers from Crepy-en-Valois.

### Comparison of the assays and estimation of the prevalence in different population subsets

Results obtained with sera from each category of individuals are presented Fig. 1. The pre-epidemic samples served as negative controls. With the four assays, signals were consistently negative (S-Flow and LIPS S1) or low (ELISAs and LIPS N). This strongly suggested that a prior exposure to human seasonal coronaviruses associated to the “common cold” (such as HCoV-OC43, HCoV-229E, HCoV-HKU-1 or HCoV-NL63) does not induce an obvious cross-reaction with our assays. This was expected, since these prevalent viruses are distantly related to SARS-CoV-2 at the protein level. With each assay, we established cut-off thresholds. For ELISA N, the cut-off was set at 95% percentile of 491 pre-epidemic sera. For ELISA tri-S, the cut-off was established as the mean + 2 standard deviations (SD) of the 100 pre-epidemic samples analyzed, which corresponds to 95% specificity. For the S-Flow, we established a cut-off that corresponded to a signal >20% of cells positive by flow cytometry. For the LIPS assays, the cut-off was based on internal controls. S-Flow and LIPS S1 cut-offs eliminated all pre-epidemic samples analyzed.

Having established these cut-off levels, we analyzed samples from 51 patients from Hôpital Bichat. Some of the patients were analyzed at different time points, representing a total of up to 161 samples. The percentage of positive samples varied between 65 and 72%, with a mean of 64%. The fact that not all patients were seropositive reflected the various sampling times from each individual. To study more precisely the kinetics of seroconversion, we selected 5 patients with more than five longitudinal samples and known dates of symptom onsets (Fig. 2). In these patients, seroconversion was detected between 5-10 days post symptom onsets with ELISA-N, LIPS-N, ELISA tri-S and S-Flow. The LIPS S1 assay became positive with a slower kinetic, and one of the patients remained just below the cut-off. For some patients, the LIPS N and ELISA N signals appeared before the LIPS S1 and ELISA tri-S, which suggest different kinetics of N- and S/S1 responses independently of the sensitivity of the test.

We then tested the 209 sera obtained from pauci-symptomatic individuals in Oise. Positivity rates varied from 27% to 36% between the assays, with a mean of 32% (Fig. 1 and Table 3). This range of variation was more marked than with hospitalized patients, likely because pauci-symptomatic COVID-19 individuals display lower viral loads than those requiring hospitalization and may generate lower levels and different patterns of antibodies. To our knowledge, these figures represent one of the first evaluations of SARS-CoV-2 prevalence in pauci-symptomatic individuals within a cluster of severe cases. The fact that only one third of the individuals were tested positive suggests that some of them may not have seroconverted at the time of sampling, and/or that other viruses or environmental causes were responsible for the reported symptoms.

**Table 3:** SARS-Cov-2 antibody detection

| Cohort | ELISA N | ELISA tri-S | S-Flow | LIPS<br>S1+ N | Antibody<br>prevalence |
| --- | --- | --- | --- | --- | --- |
| Pre-epidemic<br>individuals | 23/491<br>(5%) | 5/100<br>(5%) | 0/240<br>(0%) | 3/280<br>(1%) |  |
| Hospitalized<br>patients | 33/51<br>(65%) | 35/51<br>(69%) | 21/29<br>(72%) | 35/51<br>(69%) | 69%<br>(65-72%) |
| Pauci-symptomatic<br>individuals | 56/209<br>(27%) | 75/209<br>(36%) | 73/209<br>(35%) | 68/209<br>(32%) | 32%<br>(27-36%) |
| Blood<br>donors | 0/200<br>(0%) | 2/16 | 6/200<br>(3%) | 0/200<br>(0%) | 3 %<br>(0-3%) |

We next examined SARS-CoV-2 seroprevalence in samples collected from blood donors on March 20-24, 2020. Eligibility criteria for blood donation included an absence of recent signs of infection or antibiotic treatment. The donors can thus be considered as asymptomatic individuals with stringent criteria. The donors were negative with ELISA-N and LIPS assays. With S-Flow, 6 donors were positive, including two with a strong signal. These 6 positive and 10 negative donors were then tested with ELISA tri-S, and only the two strong responders scored positive. Therefore, the positivity rate in this cohort was low (1-3% with the two most sensitive assays). This suggests that the virus had not circulated to a large extent in a radius of 60 kilometers around the initial clusters. It is also likely that asymptomatic infection induces low and delayed seroconversion. Further studies are warranted to evaluate SARS-CoV-2 prevalence in denser population environments.

### Correlations between assays

We performed a side-by-side comparison of the assays using the three cohorts. For a given assay, we first scored the number of positive samples measured with the other assays (Fig. 3). With hospitalized patients, roughly similar numbers of positive cases were obtained with the four assays, with the exception of LIPS S1, confirming that this assay is less sensitive, probably because it does not catch antibodies targeting other S domains. However, combining the LIPS S1 and N results gave similar detection rates than any of the three other tests. With the cohort of pauci-symptomatic individuals, the S-Flow and ELISA tri-S yielded very close results and higher detection rates than the other tests. In blood donors, positive cases were only detected with these two tests.

**Fig. 3.**
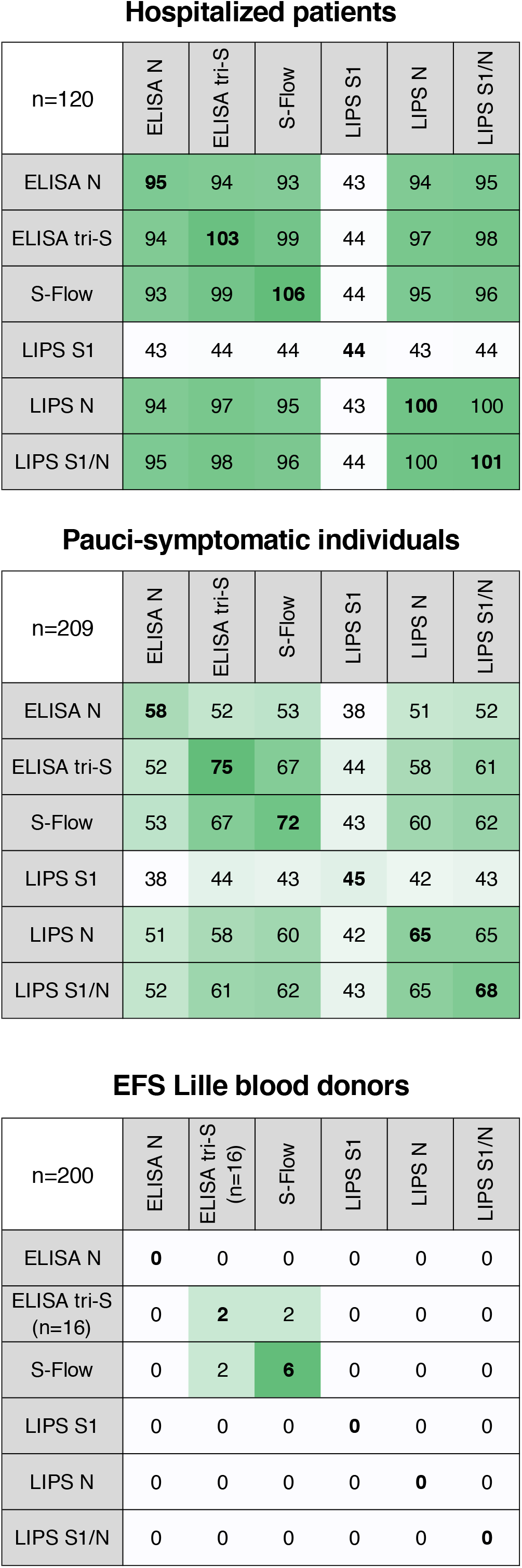
Comparison of the number of positive samples. Number of positive samples per assay and cohort is indicated, with correspondence with other tests. Each line indicates, for a given assay, the number of positive samples in common with the other assays. Bold numbers indicate the number of positive samples for a given assay. Values are color-coded, white corresponding to lower values and green the higher values.

We then mixed results obtained with the three cohorts and calculated correlation rates between each assay (Fig. 4). The dot plots indicate that sera with high antibody levels are generally caught by the four assays. Important differences are however observed with samples with a low antibody concentration, reflecting both the choice of the antigens and the intrinsic different sensitivities of the assays.

**Fig. 4.**
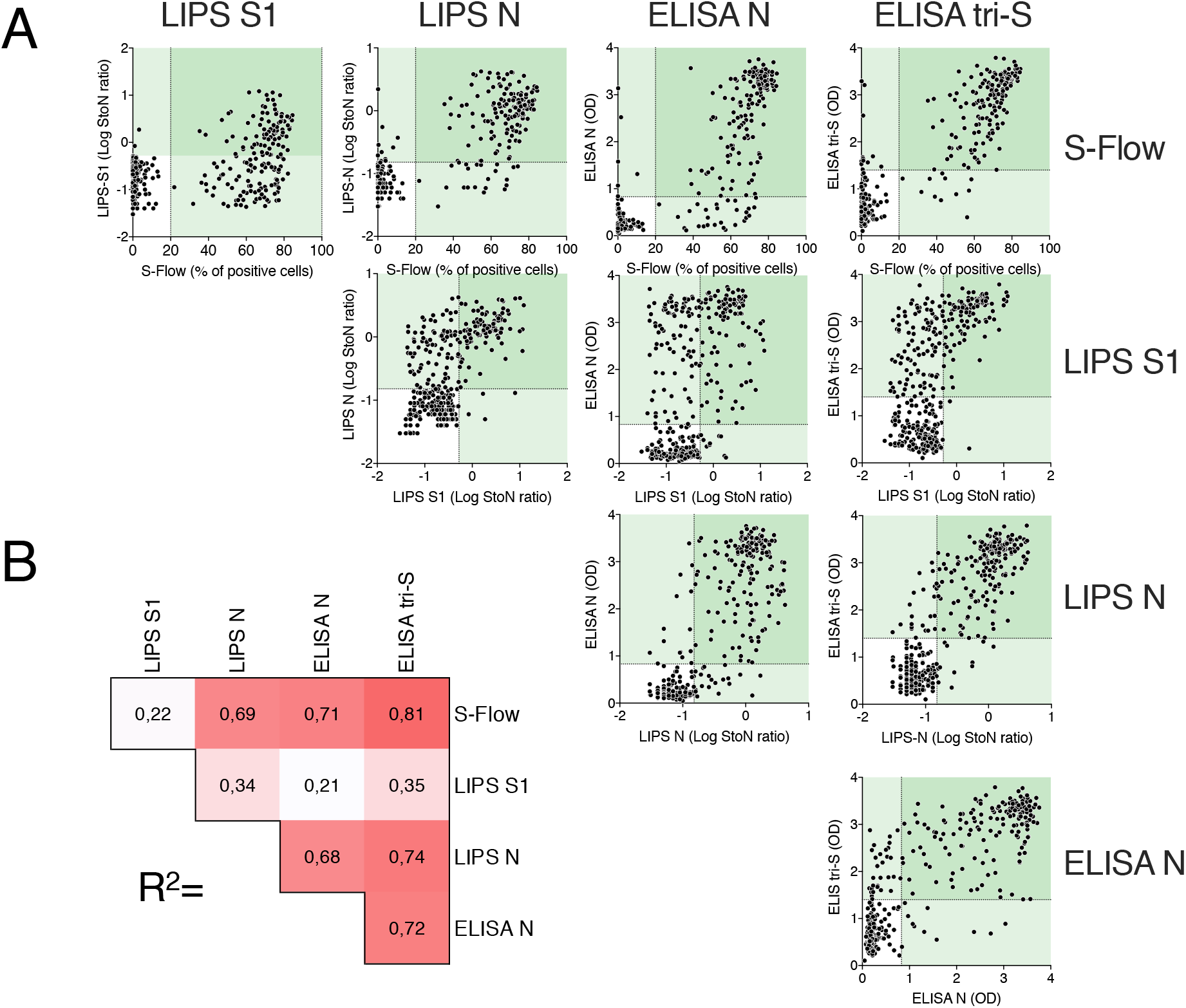
Correlations between assays. Data from pauci-symptomatic individuals and hospitalized patients (n=329) were pooled to compare assays. **A**. Data obtained with an assay were correlated to all other tests. Dashed lines indicate assays cut-offs for positivity. Values in light green areas are positive in one assay and values in dark green areas are positive in the two assays. Each dot represents a participant. **B**. Pearson correlation coefficient (R^2^) of each comparison. Values are color-coded, white corresponding to the lower value and red the highest. All correlations are significant (p>0.0001).

### Virus neutralisation assays: Microneutralisation (MNT) and Pseudovirus neutralisation

We then thought to evaluate the presence of NAbs in the sera of infected individuals. Various tests have already been established ^6,8,19,21^. We focused on two tests. The first is a microneutralisation (MNT) assay using infectious SARS-CoV-2. This reference method is based on virus incubation with serial dilutions of the sera, and evaluation of titers on Vero-E6 cells. We also developed a lentiviral-based pseudotype assay, as outlined Fig. S4A. Lentiviral particles coated with S and encoding for a reporter gene (GFP) are pre-treated with dilutions of the sera to be tested, incubated with target cells (293 T cells transiently expressing ACE2 and the TMPRSS2 protease) and the signal is measured after 48h. A pilot experiment with sera from hospitalized patients demonstrated a strong neutralizing activity with some of the samples (Fig. S4B, C). As a control, we used lentiviral particles coated with an irrelevant viral protein (VSV-G), and they were insensitive to the same sera (Fig. S4C). We also tested as a proof of concept the neutralisation activity of the first 12 sera of the cohort of pauci-symptomatic individuals (Fig. S4D). A strong correlation was observed between MNT and neutralisation of pseudoviruses (Fig. S4E). Of note, with the pseudovirus assay, similar neutralisation results were obtained when target cells transiently transfected with ACE2 and TMPRSS2 were replaced by stable 293T-ACE2 cells, or when luciferase was used as a readout instead of GFP.

The reference MNT assay is labour-intensive and requires access to a BSL3 facility. We thus performed a pilot correlative analysis between the four serological tests and the pseudovirus assay (Fig. 5A). This analysis was performed with samples from 9 hospitalized patients and 12 pauci-symptomatic individuals. A strong correlation was observed with the ELISA N, ELISA tri-S, S-Flow and LIPS-N, with a similar but less marked trend with the LIPS-S1 assay. We also determined by linear regression the association between the intensity of antibody binding and pseudovirus neutralisation. A neutralisation activity >80% was associated with the following signals: ELISA N (>2.37), ELISA tri-S (>2.9) S-Flow (>60% of positive cells) and LIPS-N (>0.049). With this level of neutralisation, LIPS S1 mainly gave positive responses and a few responses below the cut-off. In 9 hospitalized patients, the neutralisation activity increased over time, being detectable at day five and reaching 50% and 80-100% at days 7-14 and 14-21, respectively (Fig. 5B). These pilot experiments were so far performed with a limited number of samples originating from individuals with mild, severe or critical symptoms. It will be important to increase the number of pauci-symptomatic individuals tested, and to evaluate whether asymptomatic seropositive individuals exhibit a neutralisation activity.

**Fig. 5.**
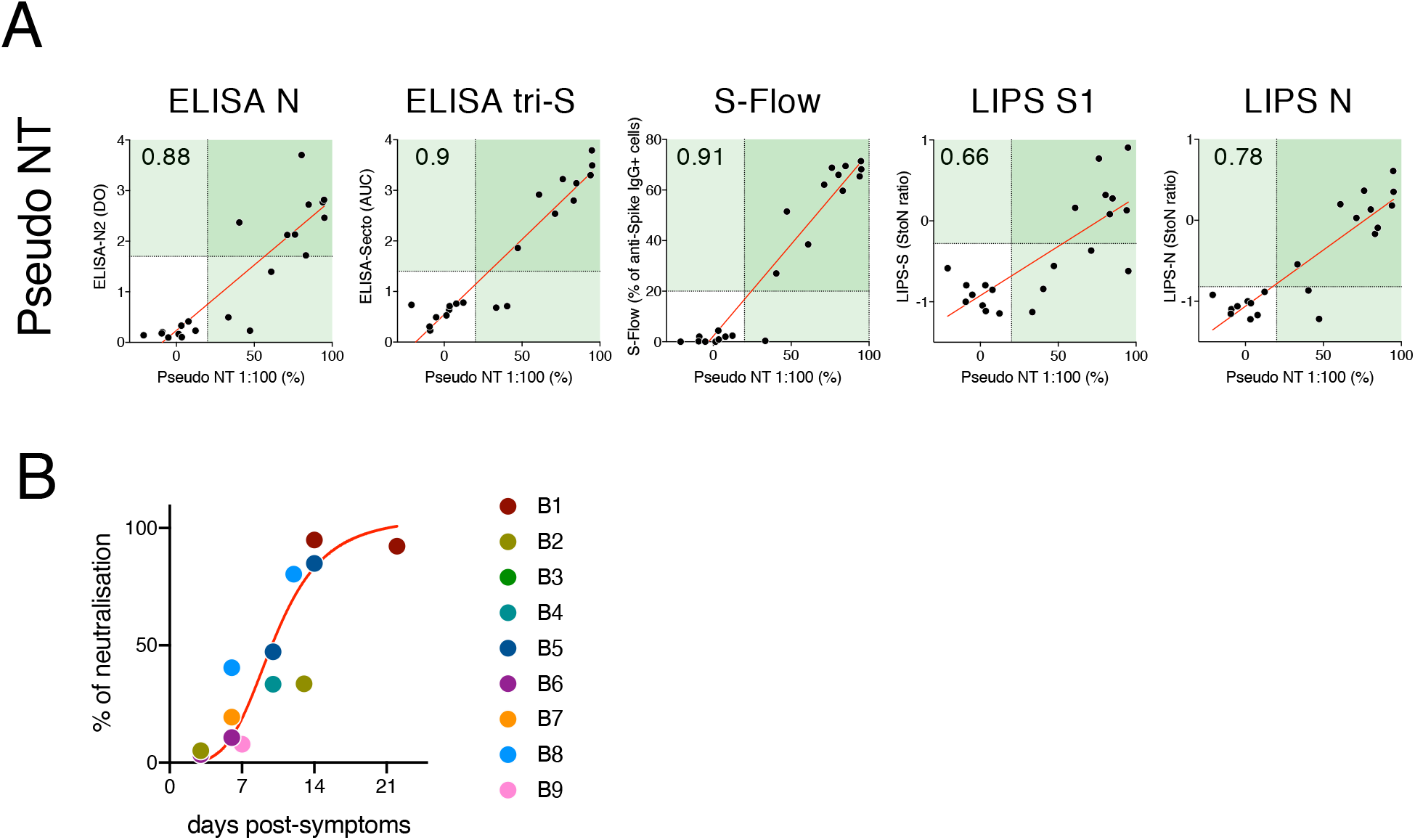
Neutralizing activity of the sera. **A**. Neutralizing activity (dilution 1:100) of 12 sera from the pauci-symptomatic cohort (C1-12) and 9 sera from hospitalized patients (B1-B9) was determined by the pseudovirus neutralisation assay and compared to serology data obtain with the 4 assays. Numbers indicate the coefficient of correlation (spearman r). All correlations are significant (p>0.0001). **B**. Neutralisation data from hospitalized patients were plotted against the day post-symptom onset. The red line corresponds to a non-linear fit of the data.

## Discussion

We have designed four serological assays to detect anti-SARS-CoV2 antibodies. The first two assays are ELISA detecting anti-N and anti-S responses. The S-Flow assay allows to identify and score the levels of antibodies binding to all domains and conformations of S expressed at the cell surface. The LIPS assays target different domains of S and N, and allow for the detailed profiling of the humoral responses. We have evaluated their performance and compared their results with two neutralisation assays, a reference MNT assay and a pseudovirus neutralisation assay.

Each assay presents advantages and drawbacks. ELISAs are widely used, either as in-house or commercial tests, and can be easily performed in routine diagnostic laboratories in large quantities. They can be performed at a high scale. The S-Flow assay captures all anti-S antibodies and provides excellent sensitivity but requires access to a cell culture system and flow cytometry equipment. Thus,

It would be less adapted to high-throughput screenings. The LIPS assay allows the testing of different target antigens in a liquid phase assay, also preserving as much as possible conformational epitopes and it appears to be as sensitive as ELISA and S-Flow for some of the antigens tested. It requires access to a bioluminescence detection instrument. The two neutralisation assays require cell culture systems, with MNT using infectious virus and necessitating access to a BSL3 facility, whereas pseudovirus neutralisation is adaptable to high-throughput screenings.

Serological diagnostic tests are complementary to viral detection by RT-PCR for diagnostic purposes in patients. Results from our study and others indicate that in severe and critical cases (hospitalized patients), seroconversion is detectable as soon as 5-14 days post symptom onset ^6,7,13-15^. In such cases, antibody titers can reach high levels, and the different assays gave similar results. Detection of anti-N and anti-full S responses demonstrated similar rates of seroconversion, whereas the S1 response was delayed. The anti-N response appeared slightly more rapidly than S/S1 responses for a given type of test, which could be of interest to develop routine diagnostics tests, if confirmed.

At the population level, serological tests are used in surveys to identify persons who have been infected. Regarding the identification of pauci-symptomatic or asymptomatic individuals, we consistently observed similar levels of seroprevalence, again with different sensitivities depending on the assay. ELISA tri-S, S-Flow and the combined LIPS S1+N gave slightly higher detection rates than ELISA-N. Combining ELISA N and S assays may also increase the sensitivity of detection. In our cohort of 209 pauci-symptomatic individuals, only a minor fraction of individuals was tested by RT-PCR (not shown). It will be useful to perform a similar analysis on individuals that have been fully characterized virologically, to further assess the serological parameters of patients with diagnosed SARS-CoV2 infection.

It has been reported that in 175 convalescent patients with mild symptoms, NAbs are detected from day 10-15 after disease onset in a large fraction of patients ^20^. The titers of NAb correlated with the titers of anti-S antibodies (targeting S, RBD, and S2 regions) ^20^. A critical question is the detection of antibodies, their neutralisation potential in asymptomatic individuals, and, more generally, the correlates of protection. In our pilot study with 200 healthy blood donors, the ELISA N and LIP S1+N assays were negative, whereas six individuals scored positive with S-Flow. When reanalyzed with the ELISA tri-S, two of the six individuals were positive. These results indicate that the most sensitive assays are required for identification of asymptomatic SARS-CoV-2 infected individuals, who will likely mount a weaker response than patients experiencing a mild or severe infection. Indeed, this should not be at the expense of specificity, as this could considerably impact the predictive value of positive results in low prevalence areas. We are currently exploring the levels of antibody responses in other contemporary cohorts to address this question.

Beyond the simple detection of individuals that have been in contact with the virus, the knowledge of immune protection (or on the contrary facilitation in case of re-infection in individuals with low antibody responses) detected with sensitive tests is key to avoid misuse of serological tests. Neutralizing antibodies have a major role in preventing reinfections for many viral diseases. A major point is the relationship between *in vivo* protection and the levels of antibody binding to the virus or neutralizing it. We compared our serological assays to MNT and pseudovirus neutralisation assay, in a limited number of individuals. We observed a strong correlation between the extent of anti-full S and even anti-N response and the neutralisation capacity of the sera. We are currently examining whether antibody levels and which viral protein best correlate with neutralisation in pauci-symptomatic or asymptomatic seropositive individuals. Answering this question will help determining whether a serological high throughput assay may serve as a surrogate to estimate the level of protection at the individual or population level. This is an important parameter to understand and model the dynamics and evolution of the epidemics and define serological tools for population control.

Non-neutralizing antibodies, or neutralizing antibodies at sub-optimal doses can also lead to Antibody-Dependent Enhancement of infection (ADE). ADE exacerbates diseases caused by feline coronavirus, MERS-CoV and SARS-CoV-1 ^25-28^. ADE might thus also play a deleterious role in COVID-19. The various techniques described here are instrumental to determine the serological status of individuals or populations and establish potential correlates of disease facilitation or protection.

## Methods

### Cohorts

Pre-epidemic sera originated from 2 pre-epidemic healthy donors’ sources: 200 sera from the Diagmicoll cohort collection of ICAReB platform^29^ approved by CPP Ile-de-France I, sampled before november 2019. 200 anonymized samples from blood donors recruited in March 2017 at the Val d’Oise sites of Etablissement Français du Sang (EFS, the French blood agency). The ICAReB platform (BRIF code n°BB-0033-00062) of Institut Pasteur collects and manages bioresources following ISO 9001 and NF S 96-900 quality standards ^29^.

COVID-19 cases were from included at Hôpital Bichat–Claude-Bernard in the French COVID-19 cohort. Some of the patients have been previously described ^24^. Each participant provided written consent to participate to the study, which was approved by the regional investigational review board (IRB; Comité de Protection des Personnes Ile-de-France VII, Paris, France) and performed according to the European guidelines and the Declaration of Helsinki.

Pauci-symptomatic individuals: On Feb 24, 2020, a patient from Crepy-en-Valois (Oise region, northern France) was admitted to a hospital in Paris with confirmed SARS-CoV-2 infection. As part of an epidemiological investigation around this case, a cluster of COVID-19 cases, based around a high school with an enrolment of 1200 pupils, was identified. On March 3-4, students at the high school, their parents, teachers and staff (administrative staff, cleaners, catering staff) were invited to participate to the investigation. A 5 mL blood sample was taken from 209 individuals who reported fever or mild respiratory symptoms (cough or dyspnea) since mid-January 2020. The median age was 18 years (interquartile range: 17-45), and 65 % were female. This study was registered with ClinicalTrials.gov (NCT04325646) and received ethical approval by the Comité de Protection des Personnes Ile de France III. Informed consent was obtained from all participants.

Samples from blood donors were collected in accordance with local ethical guidelines by Etablissement Français du Sang (EFS, Lille, France) in Clermont (Oise) on March 20 and Noyon (Oise) on March 24, both cities are located at 60 kilometers from Crepy-en-Valois.

All sera were heat-inactivated 30-60 min at 56°C, aliquoted and conserved at 4°C for short term use or frozen.

### ELISA-N

A codon-optimized nucleotide fragment encoding full length nucleoprotein was synthetized and cloned into pETM11 expression vector (EMBL). The His-tagged SARS-CoV-2 N protein was bacterially expressed in *E. coli* BL21 (DE3) and purified as a soluble dimeric protein by affinity purification using a Ni-NTA Protino column (Macherey Nagel) and gel filtration using a Hiload 16/60 superdex 200 pg column (HE Healthcare). 96-well ELISA plates were coated overnight with N in PBS (50 ng/well in 50 μl). After washing 4 times with PBS–0.1% Tween 20 (PBST), 100 µl of diluted sera (1:200) in PBST–3% milk were added and incubated 1 h at 37°C. After washing 3 times with PBST, plates were incubated with 8,000-fold diluted peroxydase-conjugated goat anti-human IgG (Southern Biotech) for 1 h. Plates were revealed by adding 100 μl of HRP chromogenic substrate (TMB, Eurobio Scientific) after 3 washing steps in PBST. After 30 min incubation, optical densities were measured at 405 nm (OD 405). OD measured at 620 nm was subtracted from values at 405 nm for each sample.

### ELISA tri-S

A codon-optimized nucleotide fragment encoding a stabilized version of the SARS-CoV-2 S ectodomain (amino acid 1 to 1208) followed by a foldon trimerization motif and tags (8xHisTag, StrepTag, and AviTag) was synthetized and cloned into pcDNA™3.1/Zeo(+) expression vector (Thermo Fisher Scientific). Trimeric S (tri-S) glycoproteins were produced by transient co-transfection of exponentially growing Freestyle™ 293-F suspension cells (Thermo Fisher Scientific, Waltham, MA) using polyethylenimine (PEI)-precipitation method as previously described ^30^. Recombinant tri-S proteins were purified by affinity chromatography using the Ni Sepharose® Excel Resin according to manufacturer’s instructions (ThermoFisher Scientific). Protein purity was evaluated by in-gel protein silver-staining using Pierce® Silver Stain kit (ThermoFisher Scientific) following SDS-PAGE in reducing and non-reducing conditions using NuPAGE™ 3-8% Tris-Acetate gels (Life Technologies). High-binding 96-well ELISA plates (Costar, Corning) were coated overnight with 125 ng/well of purified tri-S proteins in PBS. After washings with PBS–0.1% Tween 20 (PBST), plate wells were blocked with PBS–1% Tween 20–5% sucrose–3% milk powder for 2 h. After PBST washings, 1:100-diluted sera in PBST–1% BSA and 7 consecutive 1:4 dilutions were added and incubated 2 h. After PBST washings, plates were incubated with 1,000-fold diluted peroxydase-conjugated goat anti-human IgG/IgM/IgA (Immunology Jackson ImmunoReseach, 0.8 µg/ml final) for 1 h. Plates were revealed by adding 100 µl of HRP chromogenic substrate (ABTS solution, Euromedex) after PBST washings. Optical densities were measured at 405nm (OD_405nm_) following a 30 min incubation. Experiments were performed in duplicate at room temperature and using HydroSpeed™ microplate washer and Sunrise™ microplate absorbance reader (Tecan Männedorf, Switzerland). Area under the curve (AUC) values were determined by plotting the log_10_ of the dilution factor values (*x* axis) required to obtain OD_405nm_ values (*y* axis). AUC calculation and Receiving Operating Characteristics (ROC) analyses were performed using GraphPad Prism software (v8.4.1, GraphPad Prism Inc.).

### S-Flow Assay

HEK293T (referred as 293T) cells were from ATCC (ATCC^®^ CRL-3216™) and tested negative for mycoplasma. Cells were split every 2-3 days using DMEM medium supplemented with 10% fetal calf serum and 1% Penicillin streptomycin (complete medium). A codon optimized version of the SARS-Cov-2 S gene (GenBank: QHD43416.1) ^1^, was transferred into the phCMV backbone (GenBank: AJ318514), by replacing the VSV-G gene. 293T Cells were transfected with S or a control plasmid using Lipofectamine 2000 (Life technologies). One day after, transfected cells were detached using PBS-EDTA and transferred into U-bottom 96-well plates (50,000 cell/well). Cell were incubated at 4°C for 30 min with sera (1:300 dilution, unless otherwise specified) in PBS containing 0.5% BSA and 2 mM EDTA, washed with PBS, and stained using either anti-IgG AF647 (ThermoFisher) or Anti-IgM (PE by Jackson ImmunoResearch or AF488 by ThermoFisher). Cells were washed with PBS and fixed 10 min using 4% PFA. Data were acquired on an Attune Nxt instrument (Life Technologies). In less than 0.5% of the samples tested, we detected a signal in control 293T cells, likely corresponding to antibodies binding to other human surface antigens. Specific binding was calculated with the formula: 100 x (% binding on 293T-S – binding on control cells)/(100 - binding on control cells). We generated stably-expressing 293T S cells during completion of this study, which yielded similar results.

### LIPS Assay

Ten recombinant antigens were designed based on the viral genome sequence of the SARS-CoV-2 strain France/IDF0372/2020 (accession no EPI_ISL_406596) obtained from GISAID database ^31^. Five targeted different domains of S: Full S1 sub-unit (residues 1-698), N-terminal domain of S1 (S1-NTD, residues 1-305), domain connecting the S1-NTD to the RBD (S1-CD, residues 307-330 and 529-700 connected by a GGGSGG linker), Full S2 sub-unit (residues 686-1208), and S441-685. For constructs that did not contain an endogenous signal peptide (residues 1-14) *i*.*e*. S1-CD and S2 constructs, an exogenous signal peptide coming from a human kappa light chain (METDTLLLWVLLLWVPGSTG) was added to ensure efficient protein secretion into the media. Five additional recombinant antigens, targeting overlapping domains of N, were designed: Full N (residues 1-419), N-terminal domain (residues 1-209), C-terminal domain (residues 233-419), N120-419 and N111-419. The LIPS assay was designed as described ^32^ with minor modifications. Expression vectors were synthesized by GenScript Company, using as backbone the pcDNA3.1(+) plasmid, with codon usage optimized for human cells. HEK-293F cells were grown in suspension and transfected with PolyEthylenImine (PEI-25 kDa, Polyscience Inc., USA). Valproic acid (2.2 mM) was added at day 1 to boost expression. Recombinant proteins were harvested at day 3 in supernatants or crude cell lysates. Luciferase activity was quantified with a Centro XS^3^ LB 960 luminometer (Berthold Technologies, France). 10^8^ LU of antigens were engaged per reaction. S1 and C-terminal domain (residues 233-419) were selected for analysing the cohorts. To increase sensitivity, the cohorts were tested at a final dilution of 1:10 of sera.

### Microneutralisation Assay

Vero-E6 cells were seeded in 96 well plate at 2.10^4^ cells/well. The day after, 100 TCID50 of virus (strain BetaCoV/France/IDF0372/2020) were incubated with serial 2-fold dilutions of sera, starting from 1:10, in 100 µl of DMEM + TPCK 1µg/ml for 1 hour at 37°C. Mixes were then added to cells and incubated for 2 hours at 37°C. Virus/sera mixes were removed, 100µl of DMEM +1µg/ml TPCK were added, and cells incubated for 72 hours at 37°C. Virus inoculum was back titrated in each experiment. CPE reading was performed by direct observation under the microscope, and after cell coloration with crystal violet. Microneutralisation titers are expressed as the serum dilution for which 50% neutralisation is observed.

### Preparation of lentiviral pseudotypes

Pseudotyped viruses were produced by transfection of 293T cells as previously described ^33^. Briefly, cells were co-transfected with plasmids encoding for lentiviral proteins, a GFP reporter (or a luciferase reporter when specified) and the SARS-CoV-2 S plasmid, or the VSV-G plasmid as a control. Pseudotyped virions were harvested at days 2-3 post-transfection. Production efficacy was assessed by measuring infectivity or p24 concentration.

### S-Pseudotype neutralisation assay

293T Cells were transiently transfected with ACE2 and TMPRSS2 expression plasmids using Lipofectamine 2000 (Life technologies) as described above. 24h after transfection cells were detached with PBS-EDTA and seeded in Flat-bottom 96-well plates. S-pseudotypes were incubated with the sera to be tested (at 1:100 dilution, unless otherwise specified) in culture medium, incubated 10 min at RT and added on transfected cells. After 48 hours cells were detached using PBS-EDTA, fixed with 4% PFA and analyzed on an Attune Nxt flow cytometer. The frequency of GFP+ cells in each condition was determined using FlowJo v10 software and neutralisation was calculated using the formula: 100 x ((mean of replicates - mean of negative controls)/(mean positive controls - mean of negative controls)). S-pseudotypes incubated without serum and medium alone were used as positive and negative controls, respectively. 293T-cells stably expressing ACE2 were also used in this assay and yielded similar results. For luciferase-expressing pseudotypes, samples were analyzed with the EnSpire instrument (PerkinElmer).

### Data processing and analysis

Flow cytometry data were analyzed with FlowJo v10 software (TriStar). Calculations were performed using Excel 365 (Microsoft). Figures were drawn on Prism 8 (GraphPad Software). Statistical analysis were calculated using Prism 8.

## Data Availability

All data generated or analysed during this study are included in this published article (and its supplementary information files).

## Acknowledgments

We thank the patients and individuals who donated their blood, Nicoletta Casartelli for critical reading of the manuscript, Jérémy Brunet, Chantal Combredet, Valérie Najburg for help, Annette Martin for discussions, the ICAReB team for management and distribution of the samples.

## Author contribution

Conceptualization and Methodology: AF, BH, ME, HM, OS and SVW

Cohort management and sample collection: LB, LLF, QLH, DD, YY, LT, CB, MNU, GM, PM, SR, AF, BH, MAC

Serological and seroneutralisation assays: LG, ST, CP, CD, CH, FGB, JD, DP, MC, IS, RR, JB, MA, KYC, BC, FD, PS, FA, SB, VE, NE, PC, TB, ME, HM, OS and SVW

Ressources: MG, NE, SP, MB, FR, VE, PC, SVW

Data assembly and manuscript writing: TB, ME, HM, OS and SVW Funding acquisition: AF, ME, HM, OS and SVW

Supervision: AF, ME, HM, OS and SVW

All authors reviewed and approved the final version of the manuscript.

## Figure legends

**Fig. S1. SARS-CoV-2 tri-S ELISA** seroreactivity. **A**. ELISA graphs showing the IgG reactivity of sera from pre-epidemic, pauci-symptomatic (Pauci-Sympt.), and hospitalized individuals against purified tri-S proteins. The *x* axis shows the serum dilution required to obtain the values of optical density at 405 nm (OD_405nm_) indicated on the *y* axis. Mean values ± SD from intra-assay duplicates are presented. (**B**) Table comparing the % of seroreactivity between groups according to AUC value categories (see Methods). (**C**) ROC graphs comparing tri-S ELISA IgG seroreactivity between SARS-CoV-2-exposed or infected individuals and pre-epidemic controls. Table on the right indicates for each ROC analysis the sensitivity and specificity values. (**D**) Representative ELISA graphs showing the IgG, IgM and IgA reactivity against purified tri-S proteins of selected sera from SARS-CoV-2-infected subjects. Mean values ± SD from intra-assay duplicates are presented. SD, serum dilution.

**Fig. S2. S-Flow assay. A**. Schematic representation of the S-Flow assay. **B**. representative examples of S-Flow data. Anti-S IgG levels in the serum of two hospitalized patients (B1 and B2) were measured. Cells transfected with a control plasmid indicate background levels (green) and cells transfected with a S-expressing plasmid (blue) identify the specific signal. **C**. Antibody titers determined by serial dilution for 2 patients (B1 and B2). Dashed lines indicated background levels and plain line specific signal. Levels of anti-S IgG and IgM antibodies in nine hospitalized patients (B1-B9).

**Fig. S3. SARS-CoV-2 LIPS assays. A**. Schematic representation of LIPS principle: recombinant antigens are incubated with patients’ sera and immune complexes are then precipitated onto a filter plate by protein A/G-coated beads. The measure of luminescence is proportional to the initial Ab titer. **B-C**. Evaluation of the reactivity of S-based (B) or N-based (C) antigens on 34 pre-epidemic (star) and 6 epidemic (square) patients. **D**. LIPS S2 seroreactivity evaluated on 55 sera of asymptomatic individuals from the Crépy-en-Valois cluster. Left panel: relation between S2 and S1 antibody responses using an identical molar concentration. Right panel: relation between S2 antibody responses and anti-S antibody binding measured by S-Flow. Dotted lines correspond to cut-off values of each assay.

**Fig. S4. Pseudovirus neutralisation assay. A**. Schematic representation of the assay. **B**. Representative examples of neutralisation using GFP-expressing S lentiviral pseudotypes (serum dilution 1:300). VSV-G pseudotypes were used as a control. **C. and D**. Nine hospitalized patients (B1-B9) and twelve pauci-symptomatic individuals (C1-12) were analyzed. **E**. Comparison of microneutralisation and pseudoneutralisation assays. Correlations were calculated using Spearman test.

